# Surrogate methods for parameter inference in transmission dynamic models

**DOI:** 10.64898/2026.09.23.26363802

**Authors:** Philip C. Lee, Karissa Huang, Sara Y. Tartof, Isabel Rodriguez-Barraquer, Nicola F. Müller, Alejandro Schuler, Joseph A. Lewnard

**Affiliations:** Center for Computational Biology, College of Computing, Data Science, and Society, University of California, Berkeley, Berkeley, California 94720; Department of Statistics, College of Computing, Data Science, and Society, University of California, Berkeley, Berkeley, California 94720; Division of Biostatistics, School of Public Health, University of California, Berkeley, Berkeley, California 94720; Division of Epidemiology, School of Public Health, University of California, Berkeley, Berkeley, California 94720; Department of Research & Evaluation, Kaiser Permanente Southern California, Pasadena, California 91101; Department of Health Systems Science, Kaiser Permanente Bernard J. Tyson School of Medicine, Pasadena, California 91101; Department of Medicine, University of California, San Francisco, San Francisco, California; Chan Zuckerberg Biohub, San Francisco, California

## Abstract

Mechanistic models of infectious disease transmission provide a framework for understanding and predicting disease dynamics, but statistical inference is often computationally prohibitive. This challenge is especially pronounced for models with high-dimensional structures and many unknown parameters, where trajectory matching may require thousands of evaluations of an expensive likelihood function. We developed a surrogate-assisted optimization framework that uses Bayesian optimization to reduce the number of evaluations of the underlying mechanistic model required for parameter inference. We evaluated the approach using simulated fitting problems based on age-structured susceptible-exposed-infected-recovered models with 4, 8, and 16 age classes and a 210-compartment age-structured rotavirus transmission model. Applying surrogate-assisted optimization substantially reduced the number of likelihood evaluations required to identify good parameter estimates, with larger improvements as model dimensionality increased. In the rotavirus model, the surrogate Bayesian optimization approach reached the benchmark after approximately 0.7 hours, compared with approximately 9.4 hours using traditional Nelder-Mead optimization. These results demonstrate that surrogate-assisted optimization can substantially reduce the computational burden of trajectory matching for complex mechanistic transmission models, particularly for higher-dimensional inference problems.

## Introduction

Infectious disease models often incorporate high-dimensional structures to represent heterogeneities relevant to the dynamics of pathogen spread within populations. For example, ordinary differential equation (ODE) models of disease transmission may include hundreds of compartments to represent varying host characteristics such as age, exposure history, and geography [1–3]. Such elements can help models better reproduce observed disease dynamics, facilitate inference of parameters related to transmission, guide predictions of future disease burden, or aid assessments of the impact of hypothetical interventions [4]. However, solutions must be obtained through numerical methods because such models typically lack analytical solutions. The considerable computational demands associated with these numerical solutions may prohibit model-based statistical inference or render models impractical for use in real-world emergency situations where rapid results are needed [5].

Trajectory matching is a common approach to fitting transmission models. This involves defining an objective function (loss function) that takes a set of model parameters as input and returns a numerical value (loss value) quantifying the quality of the fit, with smaller values indicating better fits. The model is then fit by minimizing the objective function with respect to the model parameters. For ODE-based disease models, the objective function is typically computed as the sum of squared residuals or the negative log-likelihood (NLL) comparing model predictions to observed disease incidence over time [6,7]. Importantly, each evaluation (or “query”) of the objective function requires numerically integrating the ODE system for a given parameter set, making computation expensive for complex models. Both non-gradient and approximate-gradient optimization algorithms require numerous queries of the objective function to identify minima. Consequently, trajectory matching can be exceedingly slow for high-dimensional transmission models, particularly when many unknown parameters must be estimated.

Bayesian approaches for parameter inference based on Markov chain Monte Carlo (MCMC) sampling algorithms face similar challenges, as they often require thousands (or even millions) of model evaluations to ensure robust convergence [8].

One strategy to mitigate this computational burden is surrogate modeling, which involves training a machine learning model (referred to as the “surrogate” or “emulator”) to replicate the output of a complex mechanistic model, given input parameters and initial conditions. Once trained, the surrogate can be evaluated faster than the original model, making it an effective tool for accelerating downstream analyses. This approach has been successfully applied in a variety of life-science contexts, including systems biology (e.g., ODE and partial differential equations models of molecular pathways) and biomedical engineering (e.g., finite-element simulations of cardiovascular systems), in addition to broader uses in physics, engineering, and geosciences [9]. In infectious disease modeling, surrogates have been developed to accelerate analyses of diverse transmission models, including discrete-time susceptible-infected-recovered (SIR) models [10], agent-based models [11–16], and ODE-based compartmental models [17]. These surrogates can be broadly categorized by the type of output they were designed to predict.

Surrogates may be trained to reproduce differing classes of outputs. The first comprises surrogates trained to predict a single, scalar output (vector-to-scalar). For example, Willem et al. (2014) [16] used ensembles of symbolic regressors to predict cumulative attack rates and the timing of epidemic peaks in agent-based influenza models, and they trained additional regressors to predict incremental quality-adjusted life year impacts of varicella-zoster virus vaccination, enabling rapid sensitivity analyses of the original models. As an alternative, vector-to-vector frameworks provide a common form for reproducing higher-complexity model predictions. For example, Cameron et al. (2015) [11] used functional regression surrogates to predict age-incidence and age-prevalence curves for an agent-based malaria model, Charles et al. (2023) [12] used a recurrent neural network to predict prevalence for an agent-based malaria model, and Kurul et al. (2025) [17] used deep neural networks to predict time-series outputs from ODE-based transmission models.

Applying single-output surrogate frameworks to evaluate an objective function from a given set of parameter values can also provide a basis for statistical inference. Previous applications of surrogate modeling to ODE-based transmission models have primarily focused on relatively simple systems with few unknown parameters [17], where computational burden may not be prohibitive. In this work, we evaluated the use of surrogate modeling to accelerate model fitting for large ODE-based compartmental models. Our approach uses machine learning models to predict the loss value returned by an objective function, given an input set of model parameters. Similar to Reiker et al. (2021) [15], we integrated these surrogates within a Bayesian optimization (adaptive sampling) framework to reduce the number of objective function queries required to identify optima. We applied this approach to several large compartmental models and compared its performance against traditional Nelder-Mead optimization.

## Results

### Overview

To evaluate the surrogate modeling approach, we constructed a series of example model-fitting problems based on age-structured compartmental models. For each model, we selected a set of ground-truth parameters and simulated data under a specified probabilistic observation process. Each simulated dataset defined a negative log-likelihood (NLL) objective function. Since the maximum-likelihood estimate solution for each simulated dataset was not known *a priori*, we evaluated the NLL at the ground-truth parameters and used this value as a benchmark for optimization performance. Importantly, the ground-truth parameters are not necessarily the maximum-likelihood parameters for a particular simulated dataset because sampling variation can result in parameter values other than the ground-truth values having a higher likelihood. Nevertheless, because the maximum-likelihood solution minimizes the NLL by definition, its NLL cannot exceed the NLL evaluated at the ground-truth parameters. Thus, an optimizer that successfully approaches the maximum-likelihood solution should be able to identify parameter values yielding an NLL at least as low as the ground-truth benchmark.

For surrogate optimization, we used a Bayesian optimization scheme implemented in the scikit-optimize package [18]. Bayesian optimization has been widely used for hyperparameter tuning in machine learning, where it can efficiently identify well-performing configurations when training models is computationally expensive [19,20]. Here, we adapt the Bayesian optimization algorithm to minimize an objective function based on trajectory matching. Briefly, the algorithm begins by generating an initial set of training examples *D*, consisting of input parameter values and their corresponding NLL values. A surrogate regressor is then fit to these examples to predict NLL values at new parameter points; in this work, we used random forests as the surrogate model. The surrogate provides an approximation of the NLL surface based on previously evaluated parameter sets, which then guides subsequent evaluations, aiming to reduce the number of computationally expensive evaluations of the true NLL needed to identify the optimum. Using the trained surrogate, the algorithm proposes a new point in the parameter space that balances two distinct goals: exploration and exploitation. Exploration favors sampling in regions of the parameter space where the surrogate’s predictions are more uncertain (i.e., have higher variance), thereby improving the surrogate’s overall approximation of the objective function surface. Exploitation favors sampling in regions where the surrogate predicts lower NLL values, with the goal of identifying the NLL-minimizing parameters. Once a new point has been proposed, we evaluate the true NLL at the proposed point, add the resulting input-output pair to *D*, and repeat the process (**Fig 1**). We provide additional technical details of the algorithm in the Methods.

**Fig 1.**
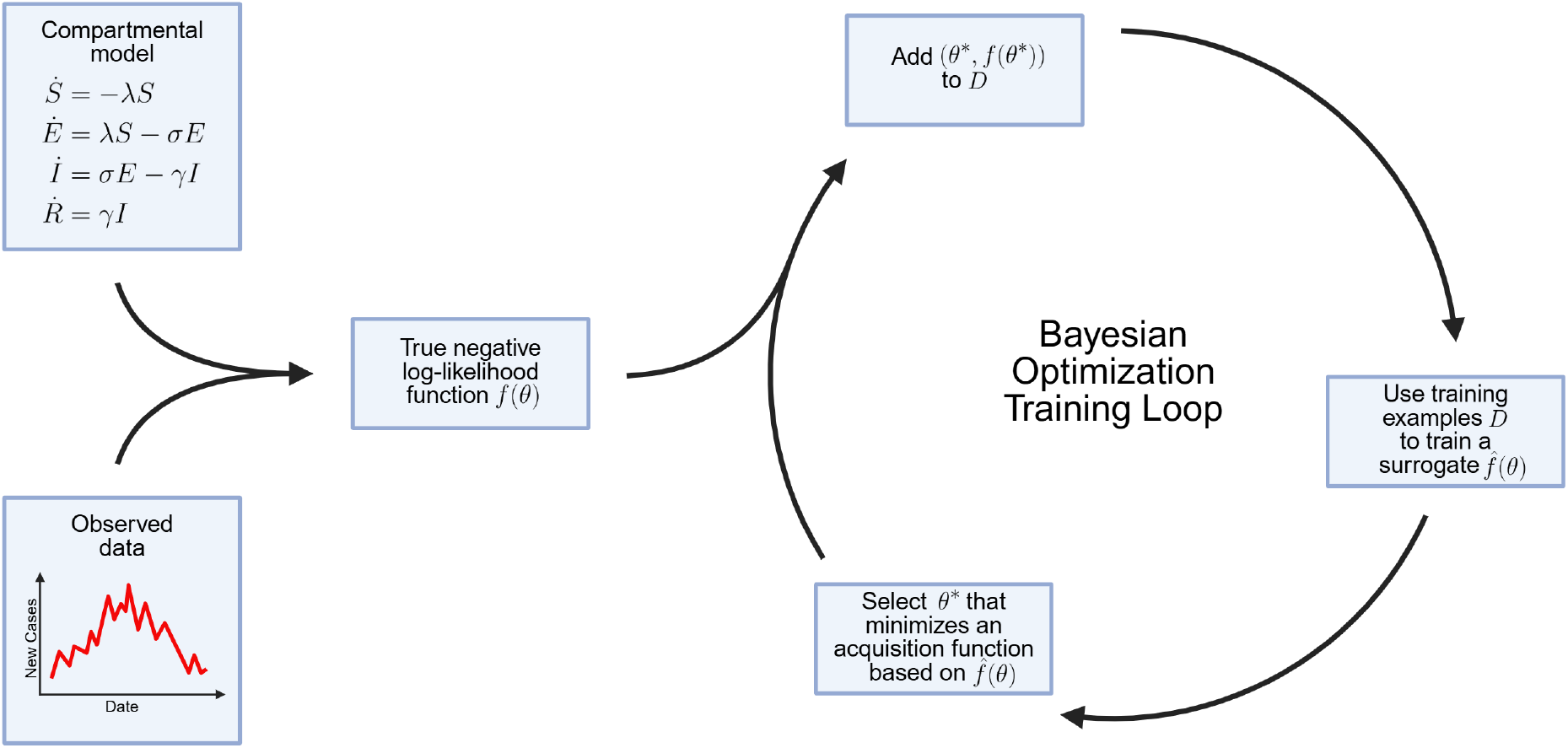
Schematic of the surrogate-based Bayesian optimization cycle. The true negative log-likelihood (NLL) function *f*(*θ*) is defined by the underlying ODE compartmental model and the probabilistic observation process linking model outputs to observed data. In the Bayesian optimization loop, a surrogate regressor 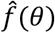 is trained using a set of training examples *D* (input parameter values and corresponding NLL values). The trained surrogate and an acquisition function are then used to propose a new point in the parameter space, balancing exploration of regions where the surrogate predictions are more uncertain with exploitation of regions where the surrogate predicts lower NLL values. The true NLL is evaluated at this proposed point, the resulting input-output pair is added to *D*, and the cycle repeats.

## Methods

We compared three optimization approaches for minimizing the NLL functions. The first was traditional Nelder-Mead optimization, a commonly used non-gradient-based method [21], which uses evaluations of the true NLL to guide the search. The second was surrogate-based Bayesian optimization, in which evaluations of the true NLL were iteratively added to the training set and used to retrain a random forest surrogate, which then guided the search. The third was a hybrid approach that combined the two strategies, beginning with surrogate-based Bayesian optimization and then switching to Nelder-Mead optimization.

### Optimization for age-structured compartmental models

We first tested the optimization approaches on NLL functions for susceptible-exposed-infected-recovered (SEIR)-type age-structured models, which were simplified versions of a model developed to study COVID-19 dynamics in British Columbia [22]. All parameters were fixed except for the age-class-specific contact-rate scaling parameters. For example, in a 16-age-class SEIR model, the simulated dataset defined a NLL function over 16 unknown scaling parameters (full model and data simulation details are provided in the **Methods)**. We generated three model-fitting problems: one with 4 age classes, one with 8 age classes, and one with 16 age classes. The most computationally expensive NLL function corresponded to the 16-age-class model, which contained 64 total compartments. We chose 16 five-year age classes as the most challenging case, as 16×16 age-specific contact matrices are commonly used to parameterize age-structured models in real-world analyses [23]. For each optimization approach, we performed 100 independent runs, each with a randomly selected initial starting point, to minimize the NLL functions for the 4-, 8-, and 16-age-class models (**Fig 2**). In these initial tests, we limited each run to a maximum of 1,000 queries (evaluations) of the true NLL function. In the hybrid approach, 500 queries were allocated to Bayesian optimization, followed by 500 queries using Nelder-Mead optimization.

**Fig 2.**
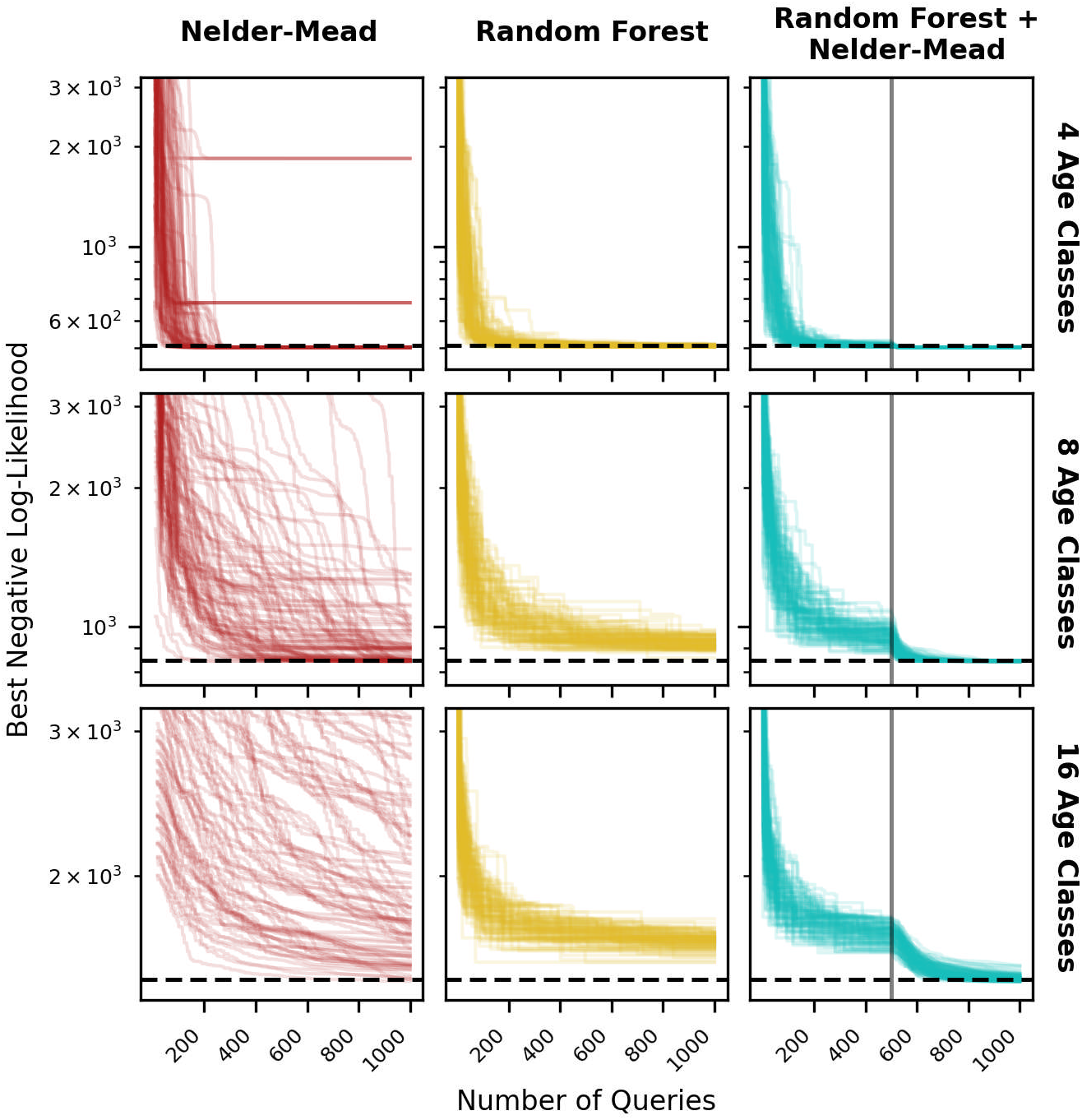
Optimization results for age-structured SEIR models. Each panel shows 100 independent optimization runs for a given combination of model (row) and algorithm (column). The three models are 4-, 8-, and 16-age-class SEIR models. The algorithms are Nelder-Mead optimization, random forest (10 trees) Bayesian optimization, and a hybrid of the two. In each panel, the horizontal dashed line indicates the negative log-likelihood (NLL) evaluated at the ground-truth parameters. The gray vertical lines in the “Random Forest + Nelder-Mead” column indicate the point at which the algorithm transitions from Bayesian optimization to Nelder-Mead optimization.

For the 4-age-class model, 88% of the Nelder-Mead runs attained NLL values at or below the ground-truth NLL; however, the remaining 12% of runs converged to NLL values very far from the ground-truth NLL (>33% away from the ground-truth NLL), likely because the optimizer became trapped in local minima (**Fig 2**). For the Bayesian optimization runs, 86% of runs attained NLL values at or below the ground-truth NLL, and the worst performing run was only around 1% away from the ground-truth NLL. The hybrid approach performed the best, where 100% of runs attained NLL values at or below the ground-truth NLL. Although the surrogate-based approaches were more consistent in minimizing the true NLL function, there was overall not a large difference among the three methods for this lower-dimensional problem.

For the 8-age-class model, the Nelder-Mead approach exhibited much greater variability than Bayesian optimization. The ground-truth NLL was 846. After 200, 500, and 1,000 queries, the median NLL across Nelder-Mead runs was 1,312 (25th-75th percentile: 1,094-2,055), 990 (25th-75th percentile: 880-1,273), and 856 (25th-75th percentile: 844-929), respectively. In contrast, the corresponding median NLLs for Bayesian optimization were 1,039 (25th-75th percentile: 989-1,111), 948 (25th-75th percentile: 925-982), and 919 (25th-75th percentile: 905-940). Bayesian optimization performed better early in the optimization, with estimated probabilities of 79%, 54%, and 29% of attaining a lower NLL than Nelder-Mead after 200, 500, and 1,000 queries, respectively. However, Nelder-Mead achieved better best-case performance: 37% of the Nelder-Mead runs attained NLL values at or below the ground-truth NLL, compared with none of the Bayesian optimization runs. The hybrid approach performed best overall, with 100% of runs attaining the ground-truth NLL, combining the early consistency of Bayesian optimization with the fine-tuning capability of Nelder-Mead.

The differences between Nelder-Mead and Bayesian optimization were even more pronounced for the 16-age-class model. The ground-truth NLL was 1,494. After 200, 500, and 1,000 queries, the median NLL across Nelder-Mean runs was 3,072 (25th-75th percentile: 2,088-4,784), 2,262 (25th-75th percentile: 1,776-3,536), and 1,848 (25th-75th percentile: 1,650-2,486), respectively. Corresponding median NLLs achieved via Bayesian optimization were 1,756 (25th-75th percentile: 1,712-1,813), 1,704 (25th-75th percentile: 1,670-1,732), and 1,665 (25th-75th percentile: 1,641-1,685). In this case, Bayesian optimization consistently outperformed Nelder-Mead, with estimated probabilities of 93%, 83%, and 72% of attaining a lower NLL than Nelder-Mead after 200, 500, and 1,000 queries, respectively. Nelder-Mead achieved better best-case performance, with 1% of runs attaining the ground-truth NLL compared with none for Bayesian optimization. The hybrid approach again performed best, with 38% of runs attaining the ground-truth NLL within the allowed 1,000 queries.

Overall, Bayesian optimization consistently performed better than Nelder-Mead early in the optimization, particularly for the higher-dimensional models, by using the surrogate to guide evaluations of the true NLL. However, the random forest surrogate was less effective at fine-tuning parameter estimates, likely because it remained a relatively coarse approximation to the true NLL surface. Switching to Nelder-Mead after an initial surrogate phase, therefore, combined the early efficiency of Bayesian optimization with the fine-tuning capability of Nelder-Mead at later stages.

We also evaluated the sensitivity of Bayesian optimization to the random forest hyperparameters (**S1 Fig**). In particular, we varied the number of trees in the random forest, sampling integers randomly between 1 and 1,000. The number of trees had little effect on the optimization performance across the SEIR models, but the training time increased substantially with the number of trees. Thus, in all subsequent analyses, we used random forests with 10 trees.

### Comparing performance with random restarts

In practice, trajectory-matching problems are often solved by repeatedly running the optimization algorithm from randomly selected initial starting points and selecting the final best-fit parameter estimates from the run that achieves the smallest NLL value—an *ad hoc* strategy aiming to mitigate risks of convergence on local minima in the loss function. Here, we use “random restart” to refer to independently rerunning the optimization algorithm from a new randomly selected initial parameter set. We compared Nelder-Mead optimization against the hybrid approach by counting the total number of NLL queries across restarts required by each method to identify parameter values with an NLL less than or equal to the ground-truth NLL. To make the comparison fair, we implemented the Nelder-Mead approach in two stages. Each run began with 500 queries of a single Nelder-Mead run. A second Nelder-Mead run was then initialized at the best parameter set identified in the first stage and allowed to continue until certain stopping criteria were met (see **Methods** for details). For the hybrid approach, the first 500 queries were allocated to Bayesian optimization, after which a Nelder-Mead run was initialized at the best surrogate-identified parameter set and allowed to proceed until the same stopping criteria were met. Implementing the purely Nelder-Mead method in two stages ensured that any differences were not due to initializing Nelder-Mead partway through the optimization process for the hybrid approach.

Using the same NLL functions as before, we performed 100 independent trials for the 4-, 8-, and 16-age-class SEIR models (**Fig 3**). Each trial consisted of repeated random restarts of the optimization algorithms until a parameter set was identified with an NLL less than or equal to the ground-truth NLL. As the number of age classes and unknown parameters increased, both approaches required more queries to attain a NLL at or below the ground-truth NLL. However, the hybrid approach was more efficient for larger models. For the 4-age-class model, Nelder-Mead required a median of 151 (25th-75th percentile: 109-195) queries, while the hybrid approach required 407 (25th-75th percentile: 291-509). For the 8-age-class model, the medians were 704 (25th-75th percentile: 599-902) for Nelder-Mead and 667 (25th-75th percentile: 632-706) for the hybrid approach. For the 16-age-class model, Nelder-Mead required a median of 5,379 (25th-75th percentile: 2,145-12,850) queries, compared to only 1,307 (25th-75th percentile: 938-1,733) for the hybrid approach.

**Fig 3.**
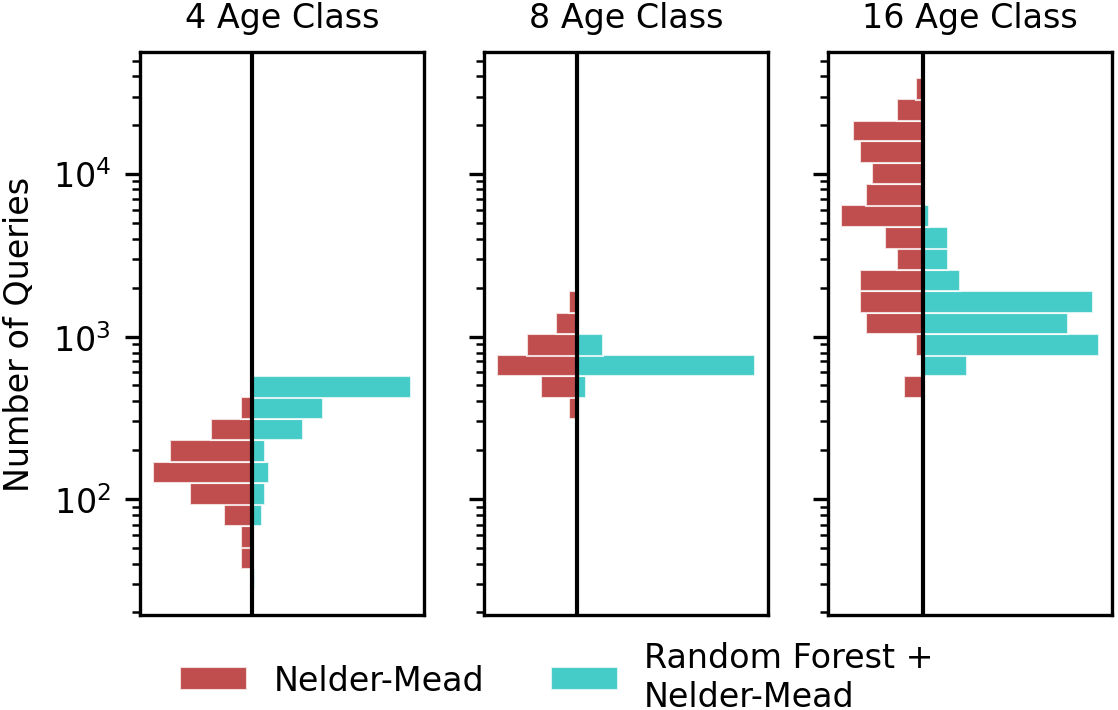
Number of queries required to reach the ground-truth negative log-likelihood (NLL) value. Each histogram shows the distribution of 100 independent trials, where each trial allowed for random restarts and terminated when parameter estimates were identified with a NLL value less than or equal to the ground-truth NLL. In the “Nelder-Mead” trials, each trial began with 500 queries of a single Nelder-Mead optimization, followed by a second Nelder-Mead run initialized at the best parameter set from the first run. In the “Random Forest + Nelder-Mead” trials, each trial began with 500 queries of surrogate Bayesian optimization before switching to Nelder-Mead optimization.

### Robustness to varying parameterizations

To evaluate robustness, we generated 100 fitting problems using different, randomly sampled parameterizations of the 16-age-class SEIR model. For each parameterization, we compared the Nelder-Mead approach with the hybrid approach, allowing for random restarts (**Fig 4**). We used the same optimization settings as before: the Nelder-Mead approach consisted of two consecutive runs, and the hybrid approach consisted of an initial phase of surrogate-based Bayesian optimization followed by Nelder-Mead optimization of the true NLL. We evaluated performance by comparing the distributions of best NLL values obtained within a fixed budget of queries of the true NLL, a pragmatic metric that approximately reflects computational burden in practical applications.

**Fig 4.**
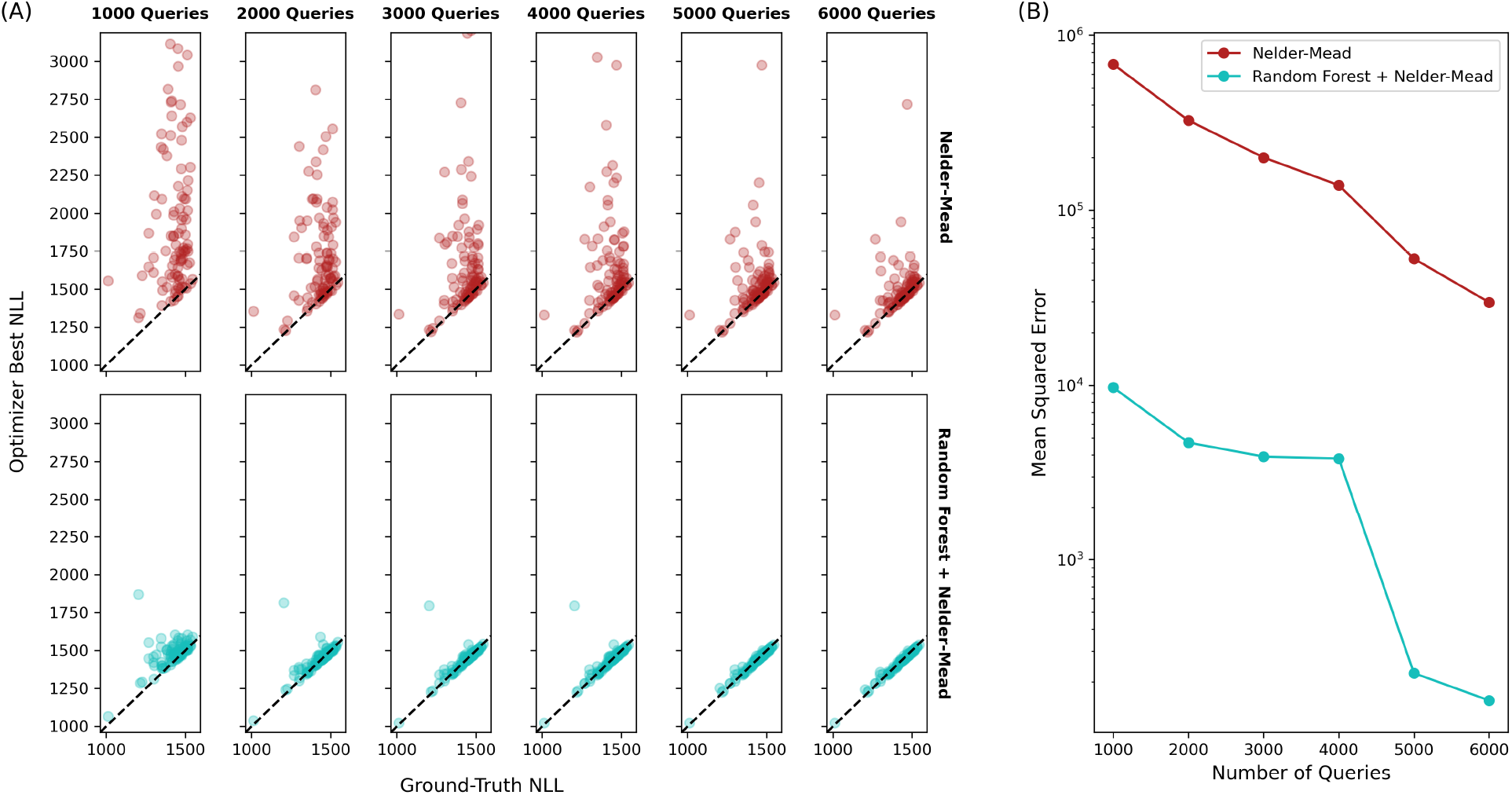
Optimization results for 16-age-class SEIR models with varying parameterizations. (A) Each point corresponds to a unique parameterization of the 16-age-class SEIR model and shows the best negative log-likelihood (NLL) identified by each optimization approach compared with the NLL at the ground-truth parameters. The top row shows results for the Nelder-Mead approach, and the bottom row shows results for the hybrid approach (random forest Bayesian optimization followed by Nelder-Mead). Columns correspond to evaluation budgets from 1,000 to 6,000 queries. The diagonal dashed lines indicate 1:1 relationships; points closer to the lines indicate NLL values closer to the ground-truth NLL. (B) Each point represents the mean squared error (MSE) calculated from the corresponding panel in (A), with the error measured relative to the ground-truth NLL for each parameterization.

For a fixed budget of NLL queries, the hybrid approach was substantially more effective at identifying parameters with NLL values close to the ground-truth NLL values. Across the 100 fitting problems, the mean squared error (MSE) between the best NLL values identified by Nelder-Mead and the corresponding ground-truth NLL values was 682,384 with a budget of 1,000 queries, whereas the MSE for the hybrid approach was 9,724. With a budget of 6,000 queries, the MSE for Nelder-Mead decreased to 29,795, while the MSE for the hybrid approach dropped to 157. Additionally, with a fixed budget of 1,000 queries, the hybrid approach identified a lower best-fit NLL than Nelder-Mead in 91/100 runs. With a budget of 6,000 queries, the hybrid approach identified a lower best-fit NLL in 82/100 runs. Thus, for the same number of queries, the hybrid approach more consistently delivered better solutions with lower NLL values.

### Optimization for rotavirus model

For a more realistic model-fitting problem, we compared the performance of Nelder-Mead and surrogate optimization on a trajectory matching problem derived from a large age-structured model previously developed to study rotavirus transmission dynamics [1]. This rotavirus model contained 210 compartments and was substantially slower to numerically integrate compared to the models evaluated in our previous theoretical examples (additional model descriptions and data simulation details are provided in the **Methods**). In the original study, the trajectory matching was performed using a NLL function that depended on four unknown parameters: the baseline transmission rate (*β*_0_), the amplitude of seasonality (*a*), the seasonal offset (*ϕ*), and the proportion of severe diarrhea cases resulting in hospitalization (*h*). Using a simulated dataset, we created a new NLL function for trajectory matching. For each optimization approach, we performed 900 independent runs with random restarts (**S2 Fig**). To simulate trajectory matching under a fixed computational time budget, we repeatedly sampled runs (with replacement) from the pool of 900 runs until the cumulative runtime of the sampled runs matched the allotted time. This procedure yielded distributions of the best NLL values identified over time for each optimization approach (**Fig 5A**). For the following results, we specifically compared the time required for the median best NLL to reach or fall below the ground-truth NLL, as well as the corresponding times for the 25th and 75th percentiles of the best NLL distribution. For example, the time at which the median best NLL reached the ground-truth NLL represents the time at which approximately 50% of optimization trials had identified an NLL at or below the ground-truth value.

**Fig 5.**
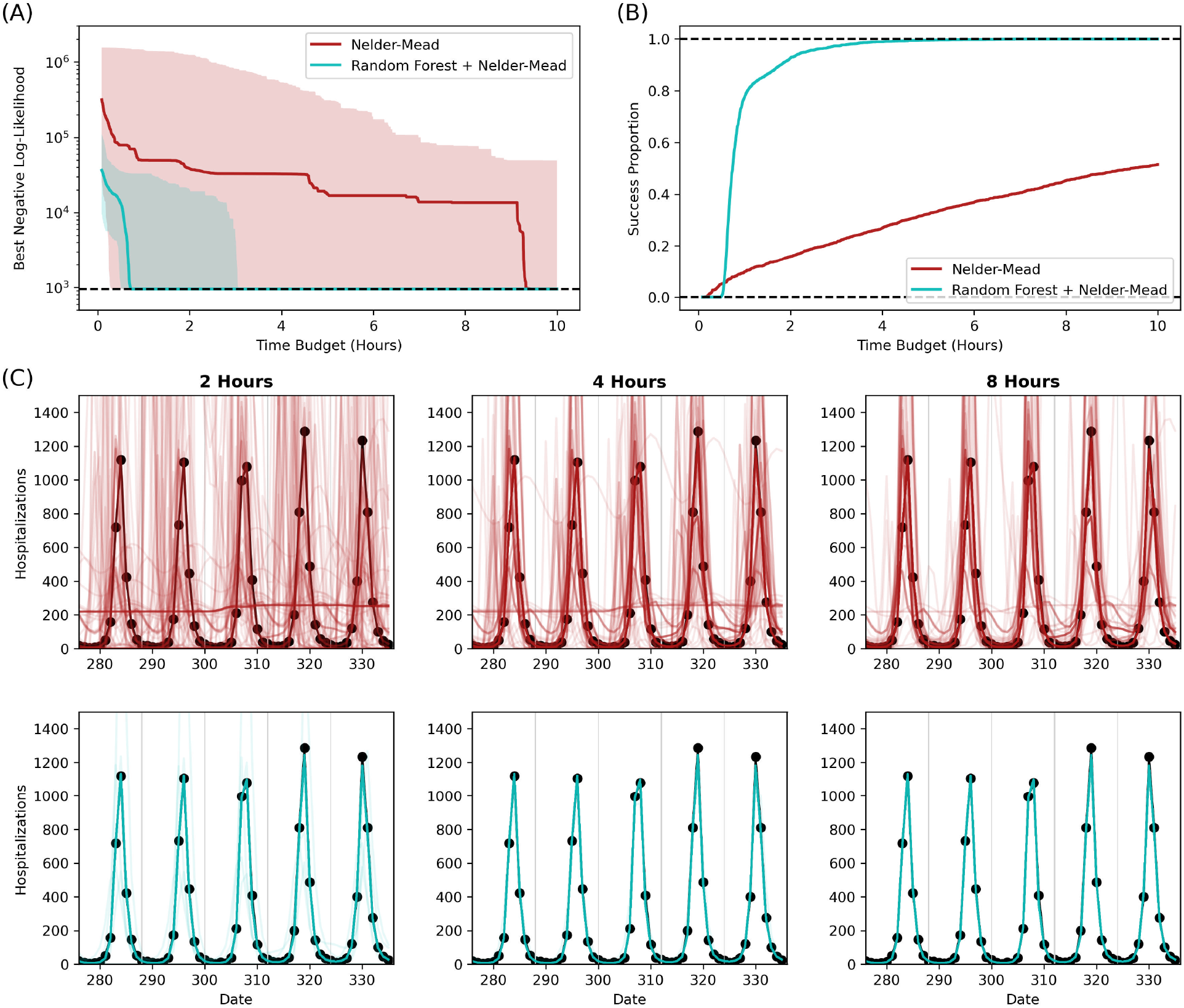
Optimization results for an age-structured rotavirus model with four unknown parameters. (A) Distributions of the best negative log-likelihood (NLL) values for Nelder-Mead and the surrogate hybrid approach over time. The distributions are based on 2,000 trials, where each trial consists of multiple runs with randomly sampled initial starting parameters. The solid lines represent median NLL values, and the shaded ribbons indicate the 2.5 and 97.5 percentiles. (B) Proportion of samples (out of the 2,000 trials) that reach the ground-truth NLL value over time. (C) Model predictions based on the best parameter estimates at different time points. The top row shows sampled model predictions from Nelder-Mead, and the bottom row shows sampled predictions from the hybrid approach. Each panel displays model predictions for 100 sampled parameter sets. Black points connected by black lines represent the simulated observed hospitalization data.

In this evaluation of a real-world problem, the surrogate hybrid approach substantially outperformed Nelder-Mead optimization. The median best NLL for the hybrid approach reached the ground-truth NLL after around 0.7 hours, with the 25th and 75th percentiles reaching the ground-truth NLL after around 0.6 and 1.0 hours, respectively. In contrast, the median best NLL for Nelder-Mead reached the ground-truth NLL after around 9.4 hours, with the 25th percentile reaching the ground-truth NLL after 3.6 hours and the 75th percentile requiring more than 10 hours. Additionally, the success proportion (proportion of trials that reached the ground-truth NLL within an allotted time budget) was overall much higher for the hybrid approach (**Fig 5B**). After 4 hours, the hybrid approach had a success proportion of 0.992, while even after 10 hours the success proportion of Nelder-Mead was only 0.524.

These differences were reflected in the quality of epidemiological trajectories generated with best-fitting parameters obtained via the hybrid and Nelder-Mead approaches, which comprised trajectories of rotavirus hospitalizations over time (**Fig 5C**). Across all time points, the distribution of model fits from the hybrid approach was more closely aligned with the simulated hospitalization data than the fits obtained using Nelder-Mead optimization.

We also constructed a more challenging variant of the original trajectory matching problem for the rotavirus model by extending the fitting procedure to address six unknown parameters. In the six-parameter problem, we added two more unknown parameters related to the relative infectiousness of second infection and asymptomatic infection—parameters fixed at sources identified in the literature in the original modeling study (additional details are provided in the **Methods**). We used the same approach to obtain distributions of the best NLL values and also the proportion of successful runs (**Fig 6, S3 Fig**). For this more challenging trajectory matching problem, the surrogate hybrid approach once again outperformed Nelder-Mead optimization.

**Fig 6.**
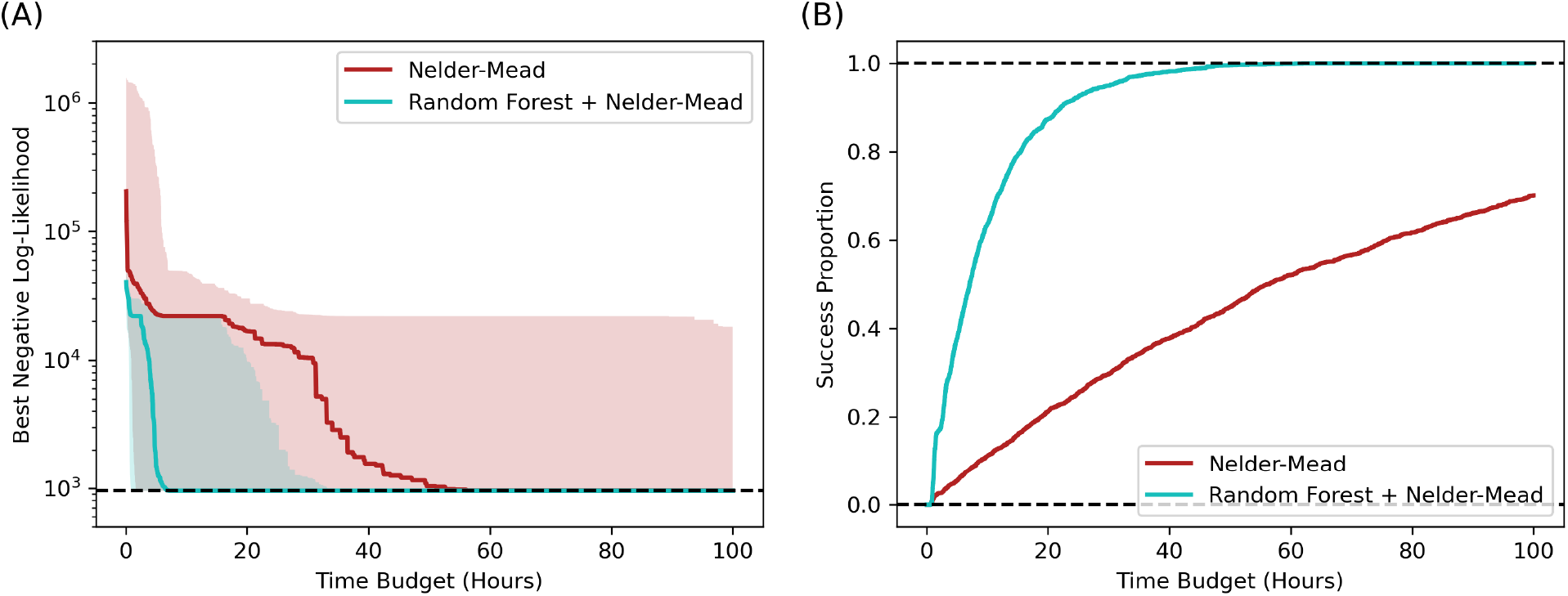
Optimization results for an age-structured rotavirus model with six unknown parameters. (A) Distributions of the best negative log-likelihood (NLL) values for Nelder-Mead and the surrogate hybrid approach over time. The distributions are based on 2,000 trials, where each trial consists of multiple runs with randomly sampled initial starting parameters. The solid lines represent median NLL values, and the shaded ribbons indicate the 2.5 and 97.5 percentiles. (B) Proportion of samples (out of the 2,000 trials) that reach the ground-truth NLL value over time.

The median best NLL for the surrogate hybrid approach reached the ground-truth NLL after around 7 hours (25th percentile: 3 hours; 75th percentile: 14 hours), while the median best NLL for Nelder-Mead reached the ground-truth NLL after around 56 hours (25th percentile: 34 hours; 75th percentile: >100 hours). After 50 hours, the success proportion of the hybrid approach was 0.997, whereas even after 100 hours the success proportion of Nelder-Mead was 0.702.

## Discussion

We investigated the use of surrogate modeling to accelerate trajectory matching for disease incidence data using ordinary differential equation (ODE)-based mechanistic models. Our approach used Bayesian optimization to construct a surrogate approximation of the negative log-likelihood (NLL) surface from previously evaluated parameter sets and used this approximation to guide subsequent evaluations of the computationally expensive true NLL. By balancing exploration of regions where surrogate predictions were uncertain with exploitation of regions predicted to have low NLL values, the approach aimed to identify good parameter estimates while reducing the number of queries of the true NLL required. Across, several representative applications, including age-structured SEIR models of increasing complexity and a large age-structured rotavirus transmission model, we found that surrogate-assisted optimization substantially reduced the number of true NLL queries required to identify optima. Moreover, we achieved substantial reductions in overall computation time when fitting the real-world rotavirus model.

Surrogate modeling had been applied previously in epidemiological contexts for two main purposes. The first was to accelerate sensitivity analyses by enabling rapid exploration of the input parameter space [16]. The second was to speed up model fitting and parameter inference [10–15,17]. These applications also differed in the form of the surrogate outputs pursued.

Jandarov et al. (2014) [10], Pokharel and Deardon (2016) [13], and Pokharel and Deardon (2022) [14], trained Gaussian process surrogates to emulate distance functions between summary statistics, which were then incorporated into approximate Bayesian computation frameworks. Reiker et al. (2021) [15] trained a Gaussian process surrogate to predict a composite goodness-of-fit score (a weighted sum of loss functions), which was then minimized to identify optimal parameters.

The choice of surrogate type is highly task-dependent, and careful consideration is needed to ensure that the computational overhead of training the surrogate does not outweigh the downstream gains in efficiency. For trajectory matching of mechanistic epidemiological models, where the goal is often to conduct statistical inference of parameter values using an objective function, our findings suggest that vector-to-scalar surrogates that directly emulate a scalar loss function can be effective. As the NLL is computed from time series outputs of incidence rates or case counts at regular intervals, vector-to-vector surrogates that predict the model time series may provide an alternative framework, with statistical inference then conducted using the predicted trajectories. However, even with a vector-to-vector surrogate that predicts the full model trajectory, parameter inference would ultimately require evaluating a scalar measure of agreement between the predicted and observed time series, such as a likelihood or loss function. Moreover, compared with vector-to-vector surrogates, vector-to-scalar models are generally faster to train because they predict simpler outputs. In theory, vector-to-vector surrogates that approximate full model trajectories may be more reusable across multiple inference tasks, whereas loss-function surrogates are inherently tied to both the mechanistic model structure and the specific observed dataset. In practice, however, trajectory matching problems are often highly context-specific. Model outputs depend not only on the governing equations but also on population characteristics such as population size and age structure. Extending vector-to-vector surrogates to account for this additional context would require expanding the input space, further increasing training time and complexity. For these reasons, we believe that vector-to-scalar surrogates represent a pragmatic and efficient choice for accelerating trajectory matching in many applied settings.

An additional advantage of our approach is that fitting of the surrogate models is embedded within a Bayesian optimization framework. Bayesian optimization is well-suited for minimizing objective functions that lack closed-form solutions and are expensive to evaluate [24]. While an ideal surrogate would be trained on a large number of examples to accurately capture the behavior of the loss function across the full extent of the parameter space, practical constraints often limit the number of available queries. Bayesian optimization addresses this limitation through adaptive sampling, also referred to as active learning, by selectively querying points that are expected to be most informative [9].

However, our surrogate modeling framework has limitations. First, vector-to-scalar surrogates that emulate the NLL are highly specific to a particular combination of mechanistic model and observed data. If new data become available or existing time-series data are extended, the surrogate-based optimization procedure must be restarted from scratch. Second, surrogate-based optimization introduces additional hyperparameters that must be selected or tuned.

Identifying suitable hyperparameter values can itself be time consuming, and care must be taken to ensure that this overhead does not offset the computational savings achieved from using the trained surrogate model. While the choice of hyperparameters for random forest models had minimal bearing on their performance in our studied examples, this cannot be assured for all applications. In practice, we recommend performing a small number of exploratory surrogate optimization runs across several reasonable hyperparameter settings and visually inspecting the best-loss-over-time curves to determine whether differences in optimization performance are large enough to warrant more extensive hyperparameter tuning.

One particularly important design choice is the selection of the surrogate model family. Although we used random forest regressors in this study, other options such as Gaussian process (GP) or neural network (NN) surrogates may be considered. Within any surrogate model family, additional hyperparameters can influence both predictive accuracy and computational efficiency. For example, Kurul et al. (2025) [17] demonstrated that the performance of NN surrogates varied substantially across different architectures. In our trajectory matching setting, we opted for random forest regressors since they were much faster to train and performance appeared to be relatively insensitive to forest size. For more challenging inference problems, GP or NN regressors, which often have higher accuracy, may be required. GP and NN regressors are substantially slower to train, and the overhead time can accumulate quickly if multiple regressors need to be repeatedly trained within a Bayesian optimization loop.

Another key hyperparameter in Bayesian optimization is the choice of acquisition function, which directs the optimization by balancing between probing areas where the loss function is likely to be small (exploitation) as well as areas where the model has higher uncertainty of the true value of the loss function (exploration). Common acquisition functions include expected improvement, where points are chosen based on how much they are expected to improve the current estimate of the optimum; lower confidence bound, where points are chosen to balance the exploration-exploitation trade-off; and probability of improvement, where points are chosen based on the probability that they improve the current estimate. In our framework, we used a combination of all three by probabilistically selecting which acquisition function to use based on historical performance (see **Methods** for additional details). Although we found that probabilistically selecting among acquisition functions performed well in our applications, the relative performance of different acquisition functions may depend on the characteristics of the optimization problem, and users applying this approach to new models or datasets should consider evaluating this sensitivity.

Under a hybrid optimization approach such as we employed, the transition point from surrogate Bayesian optimization to Nelder-Mead optimization must also be specified. In the examples that we tested, we simply picked a fixed number of queries at which the transition occurs based on the stabilization of NLL optima identified by the surrogate Bayesian optimization framework with a fixed number of iterations, typically below 500. Alternatively, an adaptive strategy could trigger the transition after a specified number of iterations without improvement. Both approaches require preliminary experimentation to identify reasonable thresholds, introducing additional overhead. In practice, a small number of exploratory runs can be used to assess when the best NLL begins to stabilize and inform selection of an appropriate transition point.

In conclusion, we introduced a surrogate modeling framework based on Bayesian optimization to accelerate statistical inference via trajectory matching in ODE-based transmission models. We showed that a hybrid approach combining surrogate Bayesian optimization with Nelder-Mead optimization can substantially improve computational efficiency in high-dimensional parameter spaces. While further gains may be achievable through more careful hyperparameter tuning, the relative insensitivity of our results to key design choices, such as random forest size, suggests that our approach is robust and practical. Future work could extend this framework to other inference tasks, including Markov chain Monte Carlo sampling and likelihood-based uncertainty quantification.

## Methods

### Compartmental model description

The SEIR-type age-structured compartmental models used in this study are simplified versions of a model developed to study COVID-19 dynamics in British Columbia [22]. In these models, individuals begin in the susceptible (*S*) compartment and, upon infection, progress through the exposed (*E*) and infected (*I*) compartments before ultimately recovering and moving to the recovered (*R*) compartment. The population is divided into *n* age classes indexed by *i* = 1, ⋯, *n*. The total population size of age class *i* is *N*_*i*_ = *S*_*i*_ + *E*_*i*_ + *I*_*i*_ + *R*_*i*_. The dynamics are governed by a transmission parameter *β* and an age-specific contact matrix *C*, where *C*_*ij*_ is the contact rate between individuals in age classes *i* and *j*. In addition, each age class includes a contact-rate scaling parameter *ϕ*_*i*_ that captures age-specific differences in susceptibility. The force of infection for a susceptible individual in age class *i* is

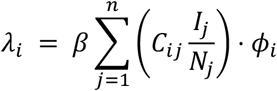

The rate of progression from *E* to *I* is *σ*, and the rate of recovery from *I* to *R* is *γ*; these rates are assumed to be identical across all age classes. Demographic processes that alter the age structure of the population are not included due to the short period evaluated. The full system of differential equations for the SEIR model is

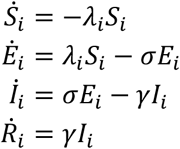

To track incident infections, we also included an auxiliary compartment *Y*_*i*_ for each age class, representing the cumulative number of infected cases, with dynamics 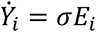. The number of new infected cases on day *t* in age class *i* is then defined as *X*_*i*_(*t*) = *Y*_*i*_(*t*) − *Y*_*i*_(*t* − 1).

### Trajectory-matching problems with SEIR model

We constructed trajectory-matching problems using SEIR models with 4, 8, and 16 age classes. To generate these problems, we first simulated observed data from each model. Parameter values were fixed at *β* = 0.1, *σ* = 1/5 days^−1^, and *γ* = 1/7 days^−1^. For the contact matrix *C*, we used the United States “All Locations” contact matrix from Prem et al. (2017), which is a 16×16 matrix binning age groups in 5-year strata (with all ages ≥80 years binned as a single category). For the 4- and 8-age-class models, age groups were aggregated and the contact matrix collapsed such that the total number of contacts between the new age groups was preserved (e.g., the first age class in the 4-age-class model corresponded to the first four age classes of the full 16-age-class matrix). The contact-rate scaling parameters *ϕ*_*i*_ were independently sampled from a uniform distribution on [0, 2]. The initial population consisted of 10,000 susceptible individuals, distributed across age classes according to the implied age distribution of the contact matrix, and a single infected individual in the first age class. The SEIR system was numerically integrated over 50 days using the odeint solver from SciPy [25], yielding the ground-truth number of new infections *X*_*i*_(t) for each age class *i* and for days *t* = 1, …, 50. Observed data *0*_*i*_(*t*) were then simulated using a Poisson observation process, where *0*_*i*_(*t*) ~ Po*i*s(*X*_*i*_(*t*)).

For trajectory matching, we assumed that *β, C, σ*, and *γ* were known, and the goal was to estimate the contact-rate scaling parameters *ϕ*_*i*_. The likelihood of observing *0*_*i*_(*t*) is *f*(*0*_*i*_(*t*); *X*_*i*_(*t; ϕ*_1_, …, *ϕ*_*n*_)), where *X*_*i*_(*t; ϕ*_1_, …, *ϕ*_*n*_) is the model-predicted number of new cases if the values of the contact-rate scaling parameters are *ϕ*_1_, …, *ϕ*_*n*_, and *f*(⋅; *λ*) is the Poisson probability mass function with mean *λ*. The joint likelihood function across all age classes and time points is

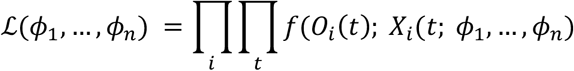

In practice, we minimized the corresponding negative log-likelihood (NLL) function, −log *ℒ*(*ϕ*_1_, …, *ϕ*_*n*_).

### Rotavirus model trajectory-matching problems

We used a large, age-structured model previously developed to study rotavirus transmission [1]. Full descriptions of this model are available in Pitzer et al. (2009) [1]. Briefly, the model consists of 11 age classes and allows for multiple infections; each infection is followed by a brief period of complete immunity, as well as an incremental reduction in susceptibility to infection and to disease upon reinfection thereafter. The model produces annual periodic patterns of rotavirus hospitalizations over many years, considering that only a fraction of all infections (depending on prior exposure history) lead to hospitalization.

To construct trajectory-matching problems, we simulated hospitalization data using the best-fit parameters from the original study, assuming a Poisson observation process. Trajectory matching was performed using the same Poisson likelihood function as in the original study. In the original formulation, the NLL function depended on four unknown parameters: the baseline transmission rate (*β*_0_), the amplitude of seasonality (*a*), the seasonal offset (*ϕ*), and the proportion of severe diarrhea cases resulting in hospitalization (*h*). We also created more challenging variants of the trajectory-matching problem involving six and eight unknown parameters. In the six-parameter problem, the relative infectiousness of secondary infection (*ρ*_2_) and asymptomatic infection (*ρ*_*A*_) were additionally treated as unknown. In the eight-parameter problem, the relative susceptibilities following first and second infections (*σ*_1_ and *σ*_2_) were additionally treated as unknown.

### Optimization algorithm

We performed Nelder-Mead optimization using the implementation in the SciPy optimize module [25] with default settings. For early stopping, we set the maximum number of function evaluations and algorithm iterations to 200 times the number of unknown parameters. Additionally, we set the values for xatol (absolute tolerance in input parameters between iterations) and fatol (absolute tolerance in function output between iterations) to 10^−4^.

We implemented Bayesian optimization using the scikit-optimize package [18]. We set the initial number of training examples to one. At each iteration, we evaluated an acquisition function at candidate parameter values to guide the selection of the next parameter value for evaluation. The acquisition function formalizes the notions of “exploration” and “exploitation” described previously by assigning a score to candidate parameter values based on the surrogate model’s predictions and uncertainty. We selected the parameter value with the lowest acquisition function value to evaluate the true NLL function. To identify the minimum of the acquisition function, we used the “sampling” strategy, in which the acquisition function was evaluated at 10,000 randomly sampled parameter values per iteration. We selected the acquisition function using the “gp_hedge” strategy, which probabilistically chooses among the lower confidence bound, negative expected improvement, and negative probability of improvement based on their historical performance. We used a random forest regressor as the surrogate model. A key hyperparameter for the random forest regressor is the number of trees. We evaluated the sensitivity of optimization performance and surrogate training time to this hyperparameter by randomly sampling integer values between 1 and 1,000 for the number of estimators (trees).

## Data Availability

All code used to produce the results and figures in this manuscript are available on GitHub at: https://github.com/pcl1701/surrogate_ml. This analysis does not involve original data.

https://github.com/pcl1701/surrogate_ml

## Code availability

All code used to produce the results and figures in this manuscript are available on GitHub at: https://github.com/pcl1701/surrogate_ml.

## Acknowledgments

The authors acknowledge support from the US Centers for Disease Control & Prevention (grant CDC-RFA-FT-23-0069 to SYT and JAL).

## Supporting information

**S1 Fig.**
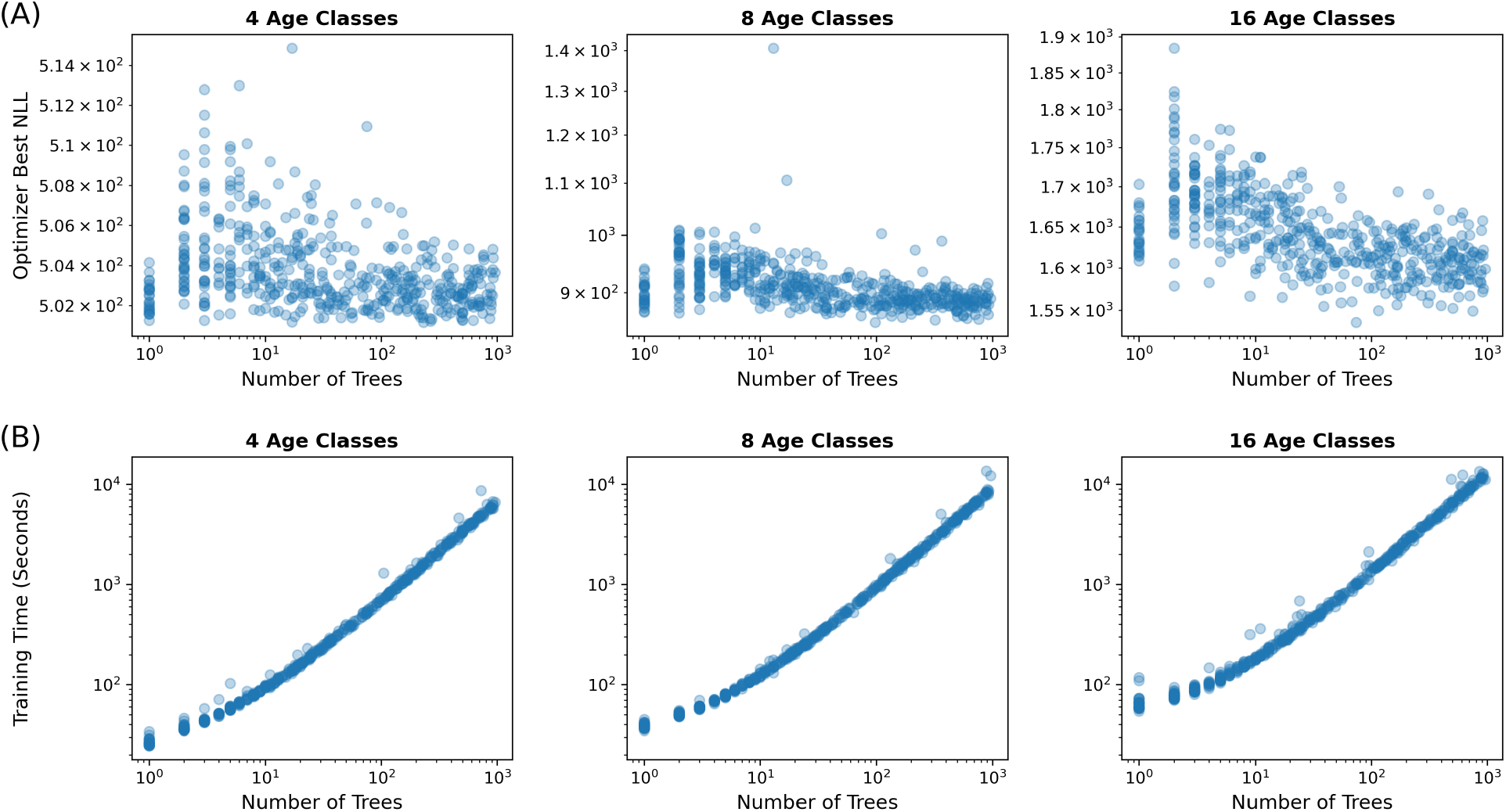
Sensitivity analysis results for random forest surrogate. (A) Best negative log-likelihood (NLL) identified using random forest surrogates in a Bayesian optimization training loop. Each point (500 points total) displays the best NLL value after 1,000 queries to the true likelihood function. (B) Total time spent in the Bayesian optimization training loop (includes time to train the random forest surrogates and also time to compute true NLL values).

**S2 Fig.**
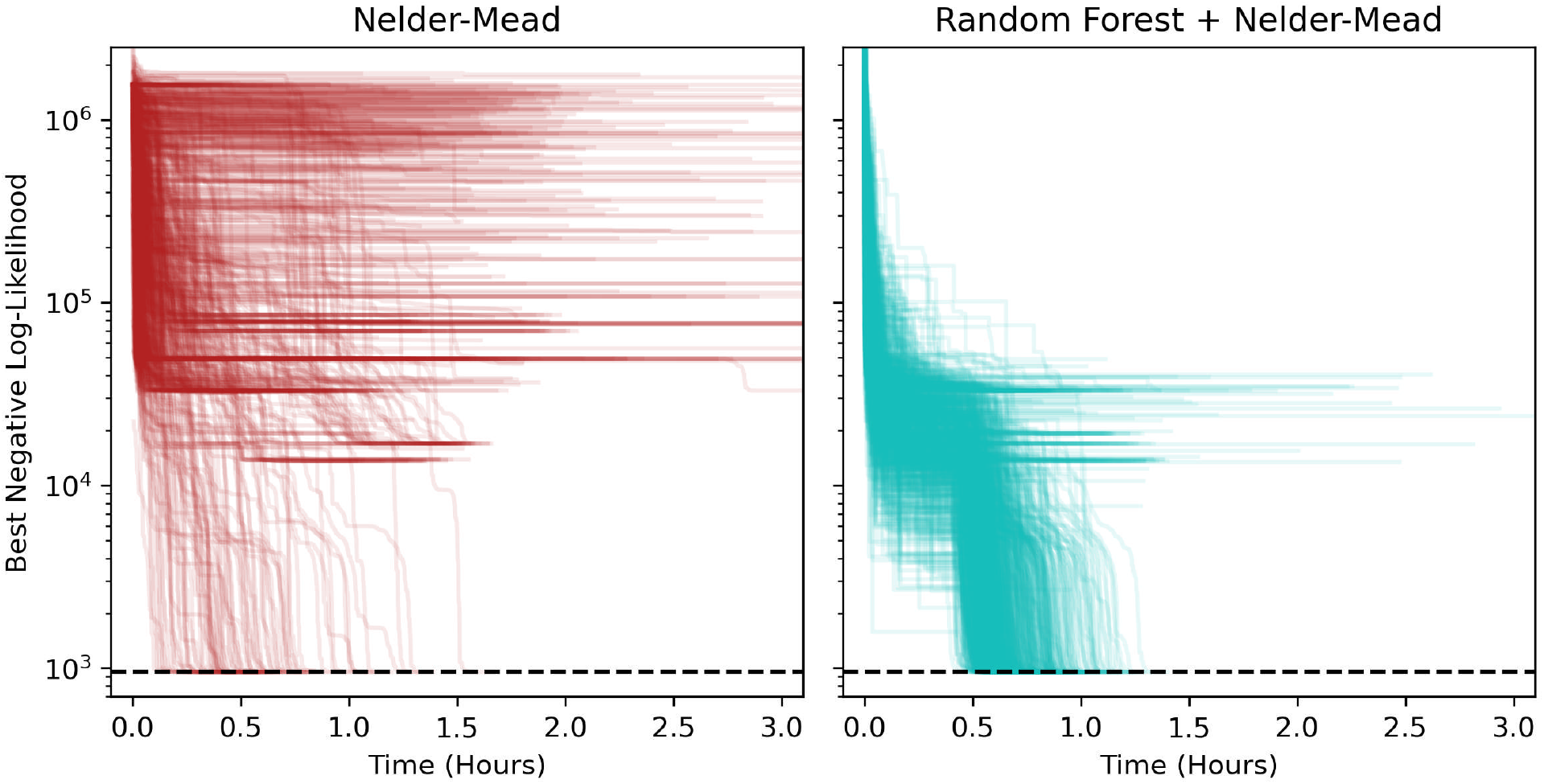
Individual runs for the 4-parameter rotavirus SIRSIRSIRS model. Each panel shows 900 independent optimization runs of the negative log-likelihood (NLL) function for the 4-parameter rotavirus trajectory matching problem. The two optimization approaches are Nelder-Mead (left) and hybrid surrogate Bayesian optimization (right). The horizontal dashed line indicates the NLL evaluated at the ground-truth parameters.

**S3 Fig.**
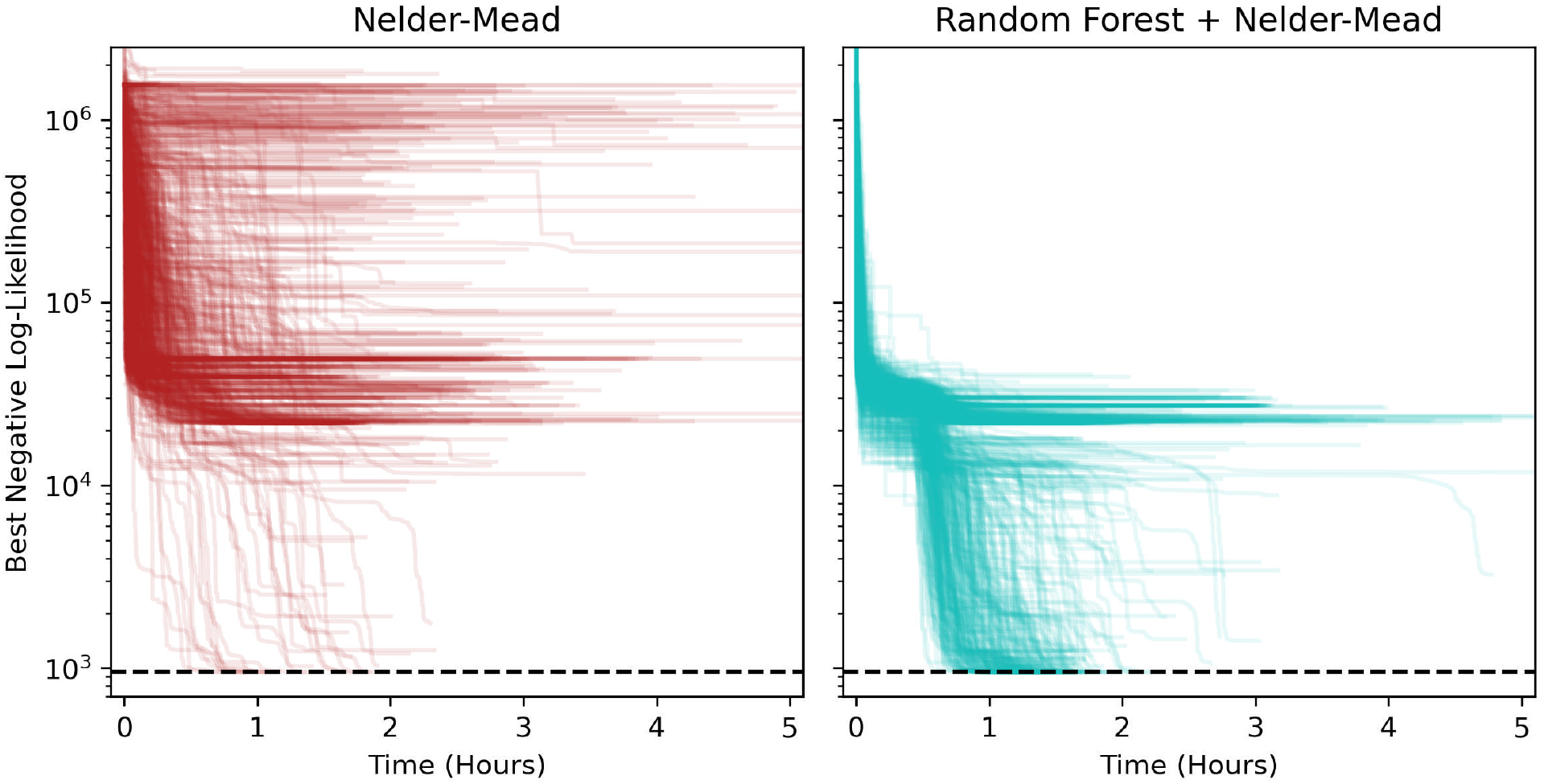
Individual runs for the 6-parameter rotavirus SIRSIRSIRS model. Each panel shows 900 independent optimization runs of the negative log-likelihood (NLL) function for the 6-parameter rotavirus trajectory matching problem. The two optimization approaches are Nelder-Mead (left) and hybrid surrogate Bayesian optimization (right). The horizontal dashed line indicates the NLL evaluated at the ground-truth parameters.

